# An evaluation of a Bayesian method to track outbreaks of known and novel influenza-like illnesses

**DOI:** 10.1101/2025.08.22.25334257

**Authors:** John M. Aronis, Ye Ye, Jessi Espino, Marian G. Michaels, Harry Hochheiser, Gregory F. Cooper

**Affiliations:** Department of Biomedical Informatics, University of Pittsburgh, Pittsburgh, Pennsylvania, USA; School of Public Health and Emergency Management, Southern University of Science and Technology, Shenzhen, China; Department of Pediatrics, University of Pittsburgh School of Medicine, UPMC Children’s Hospital of Pittsburgh, Pittsburgh, Pennsylvania, USA

## Abstract

Tracking known influenza-like illnesses, such as influenza, is an important problem in public health and clinical medicine. The problem is complicated by the clinical similarity and co-occurrence of many of these illnesses. Additionally, detecting a new or reemergent disease, such as COVID-19, is of paramount importance as recent history has shown. This paper describes the testing of a system that tracks known influenza-like illnesses and can detect the presence of a novel disease. (This manuscript is a preprint and has not been peer reviewed.)

## Introduction

Almost every year, starting in autumn and continuing through winter and into spring, a set of respiratory viruses (including influenza) emerge and circulate across the United States. The high caseloads can stress the capacity of the healthcare system, inflict suffering and high mortality rates in vulnerable populations and cost governments and healthcare systems billions of dollars each year. Additionally, there are occasional outbreaks of new or reemergent viruses such as Severe Acute Respiratory Syndrome (SARS) virus, Middle East Respiratory Syndrome (MERS) virus, and recently COVID-19. These outbreaks may be causally influenced by international travel, mutating viruses, climate change, or zoonotic transmission due to human geographic expansion.

These overlapping outbreaks of *influenza-like illnesses* (ILIs) complicate patient diagnosis and treatment. Accurately detecting and tracking these overlapping outbreaks, and recognizing the appearance of novel diseases, are critical tasks for clinical medicine and public health. By modeling known ILIs, we create a baseline with which to detect the emergence of new diseases. In addition, outbreaks of novel ILIs may begin with only a few cases distributed across a city or region that might not raise the concern of an individual clinician who may see only one or two cases near the start of the outbreak. By monitoring patient cases from across a region, we created a larger sample size with which to detect an outbreak earlier.

We previously implemented a system called *ILI Tracker* that tracks outbreaks of known ILIs in hospital emergency departments and can recognize the presence of a novel ILI. ILI Tracker starts with patient care reports (PCRs) of patients seen in emergency departments (EDs) and uses natural language processing (NLP) to extract a set of clinical findings for each patient case. It uses these findings, along with disease models built from definitive cases of known diseases, to estimate the incidence of each modeled disease in the ED population each day. ILI Tracker then analyzes whether recent patient cases are well explained by its models of known diseases, and if not, it suggests the possible presence of a novel outbreak disease in the population.

The design and implementation of ILI tracker, and the results of applying it to a small dataset containing a single year of data and a small set of findings for each patient case, are described in [1]. ILI Tracker was able to track well the incidence of a set of ILIs. It also detected an outbreak of enterovirus D68 and provided a preliminary clinical characterization of it. The current paper describes the application of ILI Tracker to a significantly larger dataset containing ten years of data and a large set of findings for each patient. We evaluate its ability to detect and track the rates of common ILIs as well as detect the initial outbreak of COVID-19, which at the time was a novel disease.

Previous approaches to outbreak detection have relied on laboratory tests, sentinel physicians, social media, and indirect indicators like medication sales, absenteeism, and news articles [2, 3, 4, 5, 6]. While laboratory data can confirm known diseases, they are ineffective against novel pathogens and are not always timely or widely available. Syndromic surveillance, which often monitors ED or other outpatient data such as chief complaints and vital signs, offers broader and timelier coverage [7, 8]. More recent systems—including time-of-arrival clustering [9], statistical term analysis [10], and semantic topic modeling [11]—have aimed to improve detection of emerging threats but still rely on narrow data scopes. The ILI Tracker system builds on prior probabilistic modeling work [12] and advances the field by leveraging a broader set of findings from patient ED reports using NLP. It explicitly models known ILIs and detects cases that do not match known diseases well as a way to detect and characterize novel diseases.

## Data and Modeling

Our data consists of records of 2,792,772 patient encounters across five emergency departments (EDs) at the University of Pittsburgh Medical Center (UPMC) from January 2011 through December 2021. UPMC serves about 58% of the residents of Allegheny County Pennsylvania, a diverse, urban and suburban county, with a population of approximately 1.2 million people We processed the data as follows:

1. We extracted coded information from each record including demographic information, laboratory tests, and other information (see below).
2. We processed each record with the NLP software MetaMap lite (version 3.6.2rc6 using the 2020AB UMLS Level 0+4+9 dataset) [13] to extract over twenty thousand findings from the narrative parts of the record. The status of each finding was recorded as being “stated as present,” “stated as absent,” or “not mentioned.”
3. We divided the data into ten outbreak years from June 1 through May 31 of each year, starting from outbreak year 2011-2012 through outbreak year 2020-2021.

Coded information includes admission date and time, demographic information (such as age, race, and gender), and vital signs (such as pulse, temperature, and blood pressure). These findings and their values are listed in Table 1. Each record also contains International Classification of Diseases (ICD) diagnosis codes relevant to the diagnosis of the ILIs we track. Additionally, each record includes results on a subset of over five thousand different laboratory results. These are described in Table 2.

**Table 1.** Demographic and physical examination findings.

| Finding | Values |
| --- | --- |
| age (years) | <5, 5-17, 18-64, >64, missing |
| gender | male, female, unknown, missing |
| ethnicity | hispanic or latino, not hispanic or latino, not specified, missing |
| race | black, white, other, not specified, missing |
| BMI | high, low, normal, missing |
| systolic blood pressure | high, low, normal, missing |
| diastolic blood pressure | high, low, normal, missing |
| pulse | high, low, normal, missing |
| temperature | high, low, normal, missing |
| respiratory rate | high, low, normal, missing |
| oxygen saturation | high, low, normal, missing |

**Table 2.** Types of laboratory tests used.

| Type | Number |
| --- | --- |
| Laboratory Chemistry and Biochemistry | 1,426 |
| Infectious Disease and Microbiology | 1,066 |
| Immunology and Allergy | 1,009 |
| Pharmacology and Toxicology | 661 |
| Hematology and Coagulation | 505 |
| Specialized Testing | 342 |
| Molecular Diagnostics and Genetics | 175 |
| Clinical Panels and Documentation | 78 |

We labeled patient ED visits as influenza cases when they either had a positive result from an influenza-specific laboratory test or included a relevant ICD code in the list of discharge diagnoses. (Laboratory tests were either PCR or ELISA based.) Those codes are listed in Tables 3 and 4. We applied this labeling criterion to six other diseases in our study to track a total of seven ILIs: influenza, respiratory syncytial virus (RSV), human metapneumovirus (hMPV), adenovirus, enterovirus, parainfluenza, and coronavirus disease 2019 (COVID-19).

**Table 3.** ICD codes related to ILIs other than influenza.

| ADENOVIRUS |  |
| --- | --- |
| 79 | Adenovirus infection in conditions classified elsewhere and of unspecified site |
| B34.0 | Adenovirus infection, unspecified |
| B30.1 | Conjunctivitis due to adenovirus |
| B30.0 | Keratoconjunctivitis due to adenovirus |
| 480 | Pneumonia due to adenovirus |
| B97.0 | Adenovirus as the cause of diseases classified elsewhere |
| COV19 |  |
| U07.1 | COVID-19 |
| ENTEROVIRUS |  |
| 8.67 | Enteritis due to enterovirus nec |
| B97.10 | Unspecified enterovirus as the cause of diseases classified elsewhere |
| B97.19 | Other enterovirus as the cause of diseases classified elsewhere |
| B34.1 | Enterovirus infection, unspecified |
| HMPV |  |
| B97.81 | Human metapneumovirus as the cause of diseases classified elsewhere |
| J21.1 | Acute bronchiolitis due to human metapneumovirus |
| J12.3 | Human metapneumovirus pneumonia |
| PARAINFLUENZA |  |
| 480.2 | Pneumonia due to parainfluenza virus |
| J12.2 | Parainfluenza virus pneumonia |
| J20.4 | Acute bronchitis due to parainfluenza virus |
| RSV |  |
| 480.1 | Pneumonia due to respiratory syncytial virus |
| 79.6 | Respiratory syncytial virus (RSV) |
| J21.0 | Acute bronchiolitis due to respiratory syncytial virus |
| J12.1 | Respiratory syncytial virus pneumonia |
| 466.11 | Acute bronchiolitis due to respiratory syncytial virus (RSV) |
| J20.5 | Acute bronchitis due to respiratory syncytial virus |
| B97.4 | Respiratory syncytial virus as the cause of diseases classified elsewhere |

**Table 4.** ICD codes related to influenza.

| INFLUENZA |  |
| --- | --- |
| J10.83 | Influenza due to other identified influenza virus with otitis media |
| 487.1 | Influenza with other respiratory manifestations |
| J11.83 | Influenza due to unidentified influenza virus with otitis media |
| 488.02 | Influenza due to identified avian influenza virus with other respiratory manifestations |
| 487.8 | Influenza with other manifestations |
| J11.81 | Influenza due to unidentified influenza virus with encephalopathy |
| J10.2 | Influenza due to other identified influenza virus with gastrointestinal manifestations |
| J10.1 | Influenza due to other identified influenza virus with other respiratory manifestations |
| J11.1 | Influenza due to unidentified influenza virus with other respiratory manifestations |
| J10.08 | Influenza due to other identified influenza virus with other specified pneumonia |
| J10.81 | Influenza due to other identified influenza virus with encephalopathy |
| J11.08 | Influenza due to unidentified influenza virus with specified pneumonia |
| J10.89 | Influenza due to other identified influenza virus with other manifestations |
| J10.00 | Influenza due to other identified influenza virus with unspecified type of pneumonia |
| 488 | Influenza due to identified avian influenza virus |
| J10.01 | Influenza due to other identified influenza virus with the same other identified influenza virus pneumonia |
| 488.01 | Influenza due to identified avian influenza virus with pneumonia |
| J10.82 | Influenza due to other identified influenza virus with myocarditis |
| J11.00 | Influenza due to unidentified influenza virus with unspecified type of pneumonia |
| J11.82 | Influenza due to unidentified influenza virus with myocarditis |
| J11.89 | Influenza due to unidentified influenza virus with other manifestations |
| J11.2 | Influenza due to unidentified influenza virus with gastrointestinal manifestations |
| 487 | Influenza with pneumonia |
| J09.X3 | Influenza due to identified novel influenza A virus with gastrointestinal manifestations |
| J09.X9 | Influenza due to identified novel influenza A virus with other manifestations |
| 488.11 | Influenza due to identified 2009 H1N1 influenza virus with pneumonia |
| J09.X1 | Influenza due to identified novel influenza A virus with pneumonia |
| 488.1 | Influenza due to identified 2009 H1N1 influenza virus |
| 488.82 | Influenza due to identified novel influenza A virus with other respiratory manifestations |
| 488.81 | Influenza due to identified novel influenza A virus with pneumonia |
| J09.X2 | Influenza due to identified novel influenza A virus with other respiratory manifestations |
| 488.89 | Influenza due to identified novel influenza A virus with other manifestations |
| 488.12 | Influenza due to identified 2009 H1N1 influenza virus with other respiratory manifestations |
| 488.19 | Influenza due to identified 2009 H1N1 influenza virus with other manifestations |

We take a Bayesian approach to patient diagnosis and outbreak detection. Thus, for each patient record with findings *E*, and each disease *D*, we model the quantity *P*(*E*|*D*), which is the probability of the patient’s clinical findings given a disease. To develop a model for disease *D*_*i*_ in testing season *S* _*j*_, we compiled a training dataset from the four seasons preceding *S* _*j*_. This dataset included all patient encounters labeled with disease *D*_*i*_. As negative training cases, we selected patient encounters from August of season *S* _*j*−1_ that did not exhibit any ICD codes or positive laboratory results that are indicative of the seven monitored ILI diseases. August was chosen based on expert consensus as a month typically exhibiting low prevalence for these diseases. An encounter in this period would have a low possibility of having any of these diseases if it did not exhibit any ICD codes or positive laboratory results consistent with those diseases. For all diseases other than those seven, we represented them as mixture disease category that we call *other*. Thus, *other* models a mixture of many diseases, including for example myocardial infarction, acute appendicitis, and pneumonia. For season *S* _*j*_, our training dataset for learning the *other* model was sourced from the cases in August of season *S* _*j*−1_. Negative cases for *other* comprised patient encounters with at least one label from the seven ILI diseases, while positive cases were those without any disease labels during August of season *S* _*j*−1_. The process described in this paragraph resulted in eight disease models (seven ILI models and a model for *other*).

For each of the seven ILI disease models we calculated information gain scores for all candidate findings from their respective training datasets. These scores measured the ability of each finding to distinguish between a diagnosed disease versus the *other* category. We selected findings as influential if their information gain scores were greater than 0·001, as determined from the training data. For a given testing season *S*_*i*_, we compiled a unified finding set by taking the union of the influential findings of all the diseases that season. We then used this comprehensive set of informative findings to model all eight diseases. (Note that the laboratory tests we used to label the disease status of training cases were not included in this set of findings.)

We constructed Naïve Bayes models featuring a binary diagnosis (*e*.*g*., influenza versus not influenza) as the parent node and the various findings as child nodes. We use the naive Bayes assumption [14] to compute the approximation:

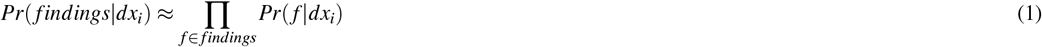

We use Laplace smoothing to avoid overreliance on rare findings:

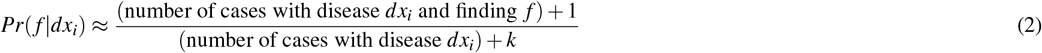

where *k* is the number of distinct values of the variable *f*. For rare findings, this quantity tends toward 1/*k*.

## Experimental Methods

Detecting the emergence of a new disease is greatly helped by first identifying and accounting for existing outbreaks of known diseases. This is especially true of ILIs which can have overlapping symptoms and similar presentations. In the work reported here, we use Naïve Bayes models to model the ILIs influenza, RSV, hMPV, adenovirus, enterovirus, parainfluenza, and COVID-19, and *other* (which includes all other diseases including, for example, acute appendicitis), for five outbreak years from 2016-2017 through 2020-2021. (Note that COVID-19 is only tracked in 2020-2021 when reliable tests and diagnoses became available.)

We derive the rate of each modeled disease using a Bayesian filter method. We start with a set of initial rates (incidences) for each modeled ILI and *other*. Then, each day, we use the rate of each disease from the previous day as a prior probability of that rate for that disease on the current day. If *Pr*(*dx*_*i*_| *f indings*) is the probability that a patient has disease *dx*_*i*_ given their *findings*, and *Pr*(*dx*_*i*_) is the prior probability of that disease in the population for the current day (the rate or posterior probability of the previous day), then by Bayes’ theorem we have the following:

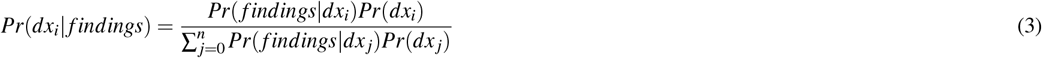

where *n* = 7, *dx*_0_ is *other*, and 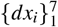 are the seven modeled ILI diseases. We can now compute the posterior probability of each disease for each patient for the current day. The expected number of patients with each disease on a given day is the sum over the probability each patient on the day has the disease. We then model the posterior probability of a given disease on that day as the expected number of patients with that disease that day divided by the total number of patients that day. On June 1 of each year, when we begin monitoring for outbreaks during that year, the initial rates of each ILI are set to low values (0·01). The ILI Tracker algorithm combines the above steps to compute the expected proportion of each disease each day: start with prior probabilities for each disease on the first day of each monitored year, compute the expected number of patients with each disease as above, then use the proportion of each disease as the prior probability of each disease on the next day. Continue this process day-by-day.

We can regard the output of ILI Tracker as a model of the types of patients who are in the ED each day. ILI Tracker assumes the presence of a fixed set of diseases that can be modeled using Naïve Bayes with specified findings. If this assumption is satisfied then the model should explain the evidence (the patients and their findings) well, and the probability of the data (the patient findings over all the patients being monitored) given the diseases in an outbreak will be relatively high.

If any of these assumptions are violated—in particular, if there are patients with a novel, unmodeled disease that presents in a clinically distinct manner from the modeled diseases—the probability of the data given the model will likely be reduced compared to a previous period of time when only modeled diseases were present in the ED. If we track the probability of the data given the output of ILI Tracker, a large decrease in the daily probability of the data may signal the presence of an unmodeled disease. An unmodeled disease may be a novel disease or a re-emergent disease that we are not currently modeling.

We can use the day-to-day patient probabilities for each disease computed by ILI Tracker to derive the likelihoods needed to monitor for a novel disease. Specifically, we can compute as follows the likelihood of the data given the output of ILI Tracker each day:

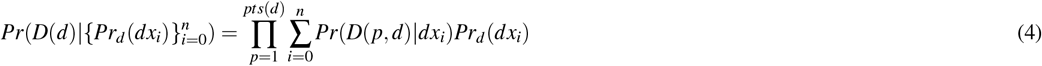

where *D*(*d*) is the data on day *d*, 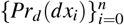 is the set of the prior probabilities of each disease on day *d* computed by ILI Tracker, *Pr*_*d*_(*dx*_*i*_) is the prior probability of disease *dx*_*i*_ on day *d*, and *D*(*p, d*) is the data for patient *p* on day *d*.

We use as a test statistic the likelihood given by Equation 4. We model the null distribution using the likelihoods for all previously monitored days within a 28-day baseline window that ended 14 days ago. We assume that this null distribution is normally distributed. We applied the Shapiro-Wilk test to a representative subset of the data during the period from June 1, 2017 to December 1, 2017, and the results of that test support the distribution is approximately normal. The *p*-value of the likelihood for the current day is the probability of that likelihood or a smaller likelihood occurring, according to the null distribution.

A complete and detailed description of this methodology is in [1].

## Results

We applied ILI Tracker to monitor the outbreak years 2016-2017 through 2020-2021. We performed four types of experiments. First, we used ILI Tracker to track known ILIs. Second, we applied ILI Tracker to evaluate if and when it detected the initial COVID-19 outbreak in Allegheny County. Third, we evaluate how ILI Tracker well detects artificial outbreaks. Finally, we evaluated the extent to which ILI Tracker could detect an outbreak of influenza when it was not provided to ILI Tracker as a modeled disease; we performed a similar experiment using RSV.

### Tracking Known ILIs

We ran ILI Tracker for each of the modeled ILIs for the outbreak years 2016-2017 through 2020-2021, and compared the daily number of labeled ED cases for each modeled disease to the daily number of ED cases of that disease predicted by ILI Tracker. Figure 1 shows ILI Tracker’s estimated daily number of cases of influenza for outbreak year 2018-2019. The strong visual correlation is supported by a Pearson *r* value of 0·69 (*p* < 0·001). Figure 2 shows ILI Tracker’s estimated daily number of cases of RSV for the same outbreak year which has a Pearson *r* value of 0·33 (*p* < 0·001).

**Figure 1.**
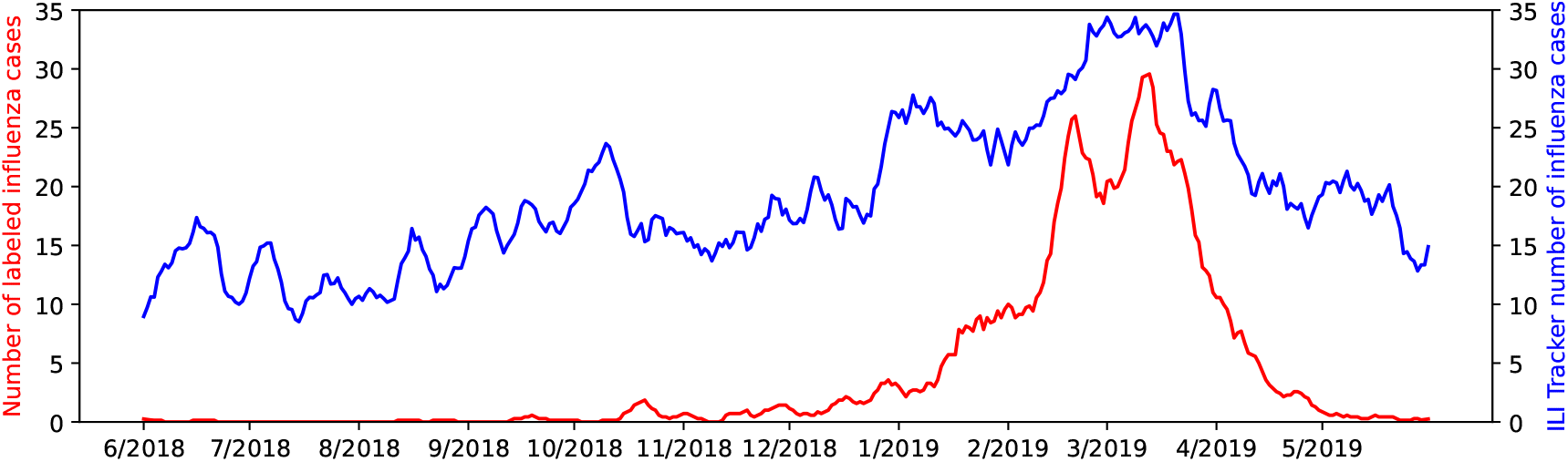
Influenza tracking during 2018-2019.

**Figure 2.**
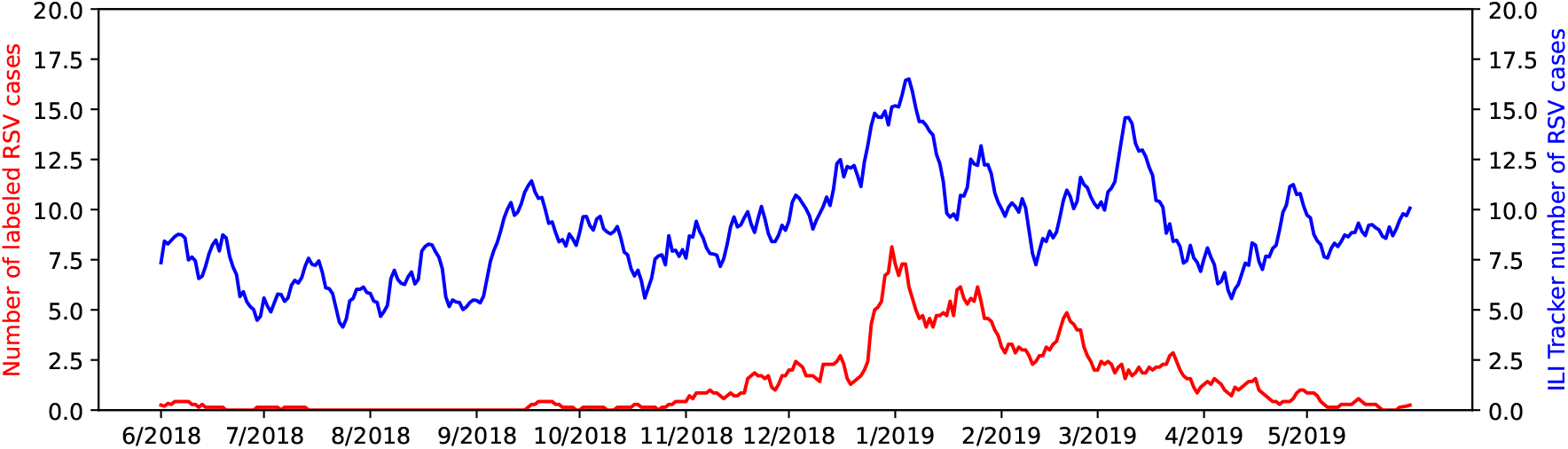
RSV tracking during 2018-2019.

Correlations for all of the modeled ILIs for each of the monitored outbreak years are shown in Table 5. The null hypothesis is that there is no correlation. Note that Pearson correlation assumes data are normally distributed and measures the strength of a linear relationship between variables. Spearman correlation does not assume data are normally distributed and measures the strength of a monotonic relationship between variables even if it is not linear, and thus it is more robust in the presence of outliers. We did not begin tracking COVID-19 until outbreak year 2020-2021 when there was sufficient data to model it.

**Table 5.** Daily correlation of ILI Tracker output with number of labeled ILIs.

| Disease | Pearson $r$ ($p$) | Spearman $r$ ($p$) | Total Labeled |
| --- | --- | --- | --- |
| 2016-2017 |  |  |  |
| influenza | 0.68 (< 0.001) | 0.65 (< 0.001) | 1,043 |
| RSV | 0.39 (< 0.001) | 0.48 (< 0.001) | 373 |
| hMPV | 0.20 (< 0.001) | 0.16 (< 0.001) | 44 |
| enterovirus | 0.04 (0.50) | 0.02 (0.71) | 188 |
| adenovirus | 0.04 (0.45) | 0.00 (0.98) | 44 |
| parainfluenza | -0.04 (0.42) | -0.04 (0.39) | 13 |
| 2017-2018 |  |  |  |
| influenza | 0.66 (< 0.001) | 0.47 (< 0.001) | 2,360 |
| RSV | 0.33 (< 0.001) | 0.40 (< 0.001) | 402 |
| hMPV | 0.06 (0.24) | 0.03 (0.61) | 82 |
| enterovirus | 0.13 (0.01) | 0.11 (0.03) | 207 |
| adenovirus | 0.10 (0.07) | 0.04 (0.46) | 40 |
| parainfluenza | 0.02 (0.69) | 0.01 (0.90) | 17 |
| 2018-2019 |  |  |  |
| influenza | 0.69 (< 0.001) | 0.65 (< 0.001) | 1,562 |
| RSV | 0.51 (< 0.001) | 0.47 (< 0.001) | 492 |
| hMPV | 0.17 (< 0.001) | 0.19 (< 0.001) | 57 |
| enterovirus | -0.01 (0.83) | -0.03 (0.61) | 433 |
| adenovirus | 0.06 (0.26) | 0.05 (0.34) | 47 |
| parainfluenza | -0.01 (0.78) | 0.01 (0.83) | 23 |
| 2019-2020 |  |  |  |
| influenza | 0.73 (< 0.001) | 0.70 (< 0.001) | 2,401 |
| RSV | 0.56 (< 0.001) | 0.66 (< 0.001) | 845 |
| hMPV | 0.12 (0.02) | 0.05 (0.30) | 48 |
| enterovirus | -0.10 (0.06) | 0.03 (0.51) | 318 |
| adenovirus | 0.08 (0.13) | 0.03 (0.51) | 67 |
| parainfluenza | 0.03 (0.61) | 0.00 (1.00) | 21 |
| 2020-2021 |  |  |  |
| influenza | 0.05 (0.36) | 0.05 (0.30) | 47 |
| RSV | 0.12 (0.02) | 0.05 (0.36) | 22 |
| hMPV | 0.02 (0.72) | 0.04 (0.50) | 1 |
| enterovirus | 0.01 (0.81) | 0.07 (0.17) | 29 |
| adenovirus | -0.02 (0.66) | -0.04 (0.48) | 4 |
| parainfluenza | 0.00 (0.94) | 0.00 (0.96) | 1 |
| COV | 0.36 (< 0.001) | 0.31 (< 0.001) | 4,902 |

### Detecting COVID-19

COVID-19 appeared in the area covered by our data sometime in the first six months of 2020, which is included in outbreak year 2019-2020, and continued through outbreak year 2020-2021 and beyond.

Figures 3 and 4 show the daily number of labeled COVID-19 patients and the *p*-values for outbreak years 2019-2020 and 2020-2021, respectively. The dashed horizontal lines indicate *p* = 0·01 (log *p* = −2, all logarithms are base 10) for reference. Note the low *p*-values in early January and late March of 2020. The low values in early January occur well before the laboratory diagnosis of COVID-19 in these EDs and may correspond to early COVID-19 patients who visited the ED. Other low *p*-values occur later in 2020 and in 2021, possibly due to outbreaks of new strains of COVID-19 that include some changes in clinical characteristics.

**Figure 3.**
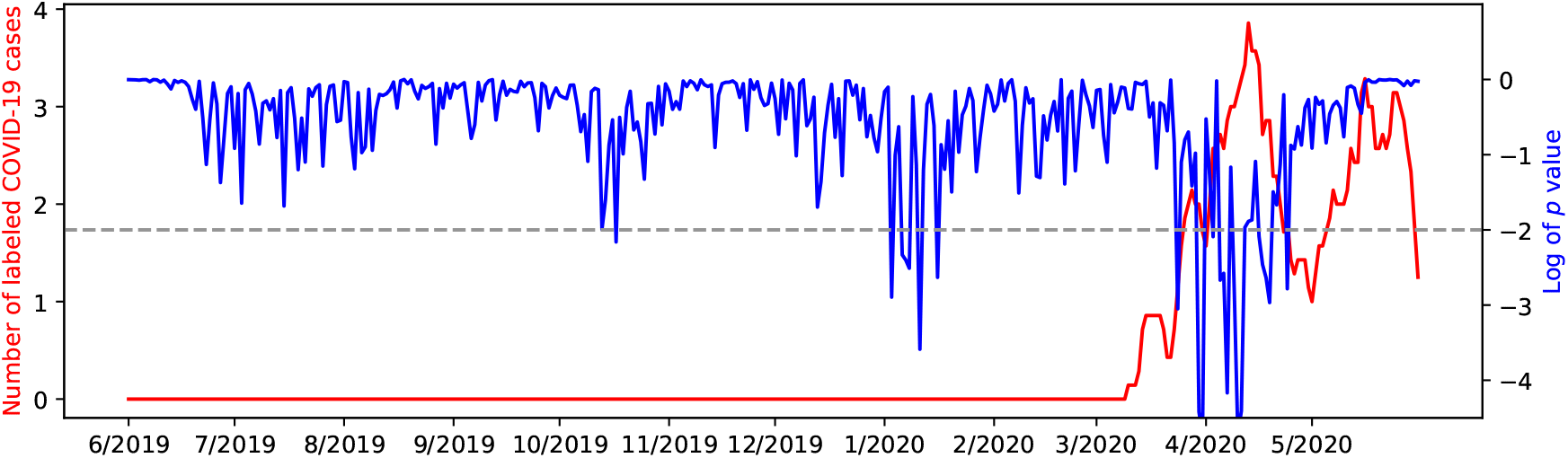
Detecting COVID-19 2019-2020.

**Figure 4.**
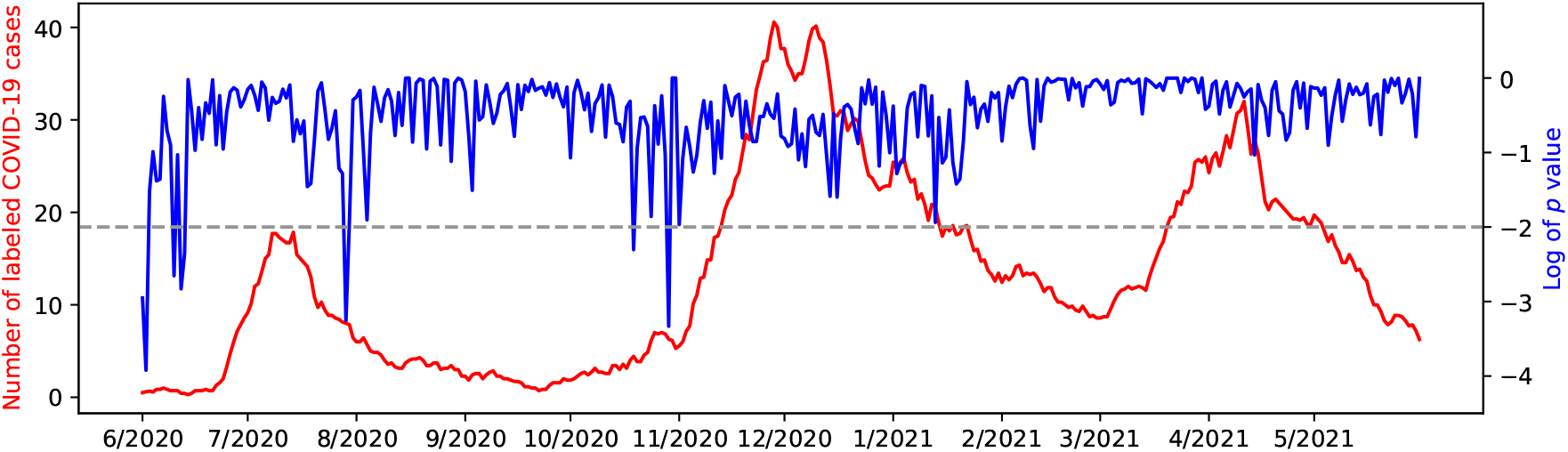
Detecting COVID-19 2020-2021.

### Detecting Artificial Outbreaks

Figure 5 shows the result of artificially adding COVID-19 patients to data from outbreak year 2017-2018. Specifically, records from 100 COVID-19 patients were added each day for the two week periods starting on September 9, December 18, and March 28 (days 100, 200, and 300). These COVID-19 cases were randomly selected from cases in outbreak year 2020-2021 that had a positive COVID-19 laboratory test result. Note the strong decrease in *p*-values during these periods. In each case, the *p*-value dropped to less than 0·01 within two days of the addition of COVID-19 patients to the data.

**Figure 5.**
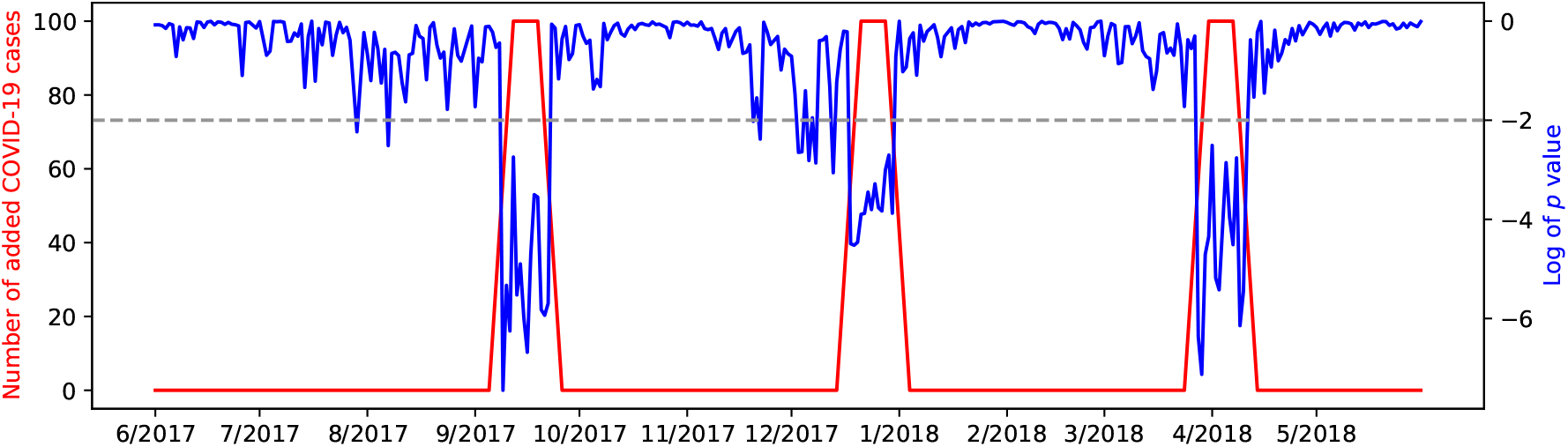
Detecting artificial outbreaks in 2017-2018.

### Leave-One-Out Disease Detection

The *p*-values computed by ILI Tracker provide a daily monitor of ED patient cases. A low *p*-value (compared to previous values) may signal the presence of a novel disease. Instead of monitoring the *p*-value every day, clinicians and public health officials may prefer an *alert* when the *p*-value is less than a predetermined *threshold* that signals the possible presence of a novel disease.

To our knowledge, there was just one major, novel outbreak in our data, namely COVID-19. To more fully test ILI Tracker we can pretend that known diseases are novel. That is, we run ILI Tracker with models of all but one known disease *dx*. We do not retain a model of *dx*, but do include models of the other known ILIs and *other*. Thus, the disease *dx* that is left out is effectively a novel disease.

We performed this experiment for influenza and RSV, for five outbreak years, for a total of ten *leave-one-out* experiments. Quantifying the results requires addressing two tasks. First, we need a way to determine the beginning of an outbreak. Second, we require a way to determine the effect of various thresholds on *p*-values. We define the beginning of an outbreak of a disease the following way:

1. We compute a Poisson distribution of the frequency of the left-out disease during the period from June 1 through August 31 when an outbreak is unlikely.
2. For each day starting with September 1, we find the *p*-value of seeing that number of labeled cases or more of the disease for that day according to the Poisson distribution.
3. We designate that an outbreak has started the first day this probability is less than 0·01.

For a particular threshold *t*, if *p* < *t* on day *d*, we say an outbreak is occurring on day *d*. The lower the *p*-value the greater the chance of an unmodeled disease.

We performed this experiment for influenza and RSV with fifty threshold values ranging from 0·001 to 0·2. For each experiment we counted the number of false alerts, defined to be alerts after September 1 but before the determined outbreak of the disease in question. We also counted the *days to detection* which is the number of days from the determined outbreak of the disease to an alert. If ILI Tracker failed to detect an outbreak we set the days to detection to sixty days. This occurred six times (out of five hundred disease-threshold combinations) for thresholds of < 0·006. Figure 6 shows the rates of false alerts and days to detection, averaged over the ten experiments, for the tested thresholds.

**Figure 6.**
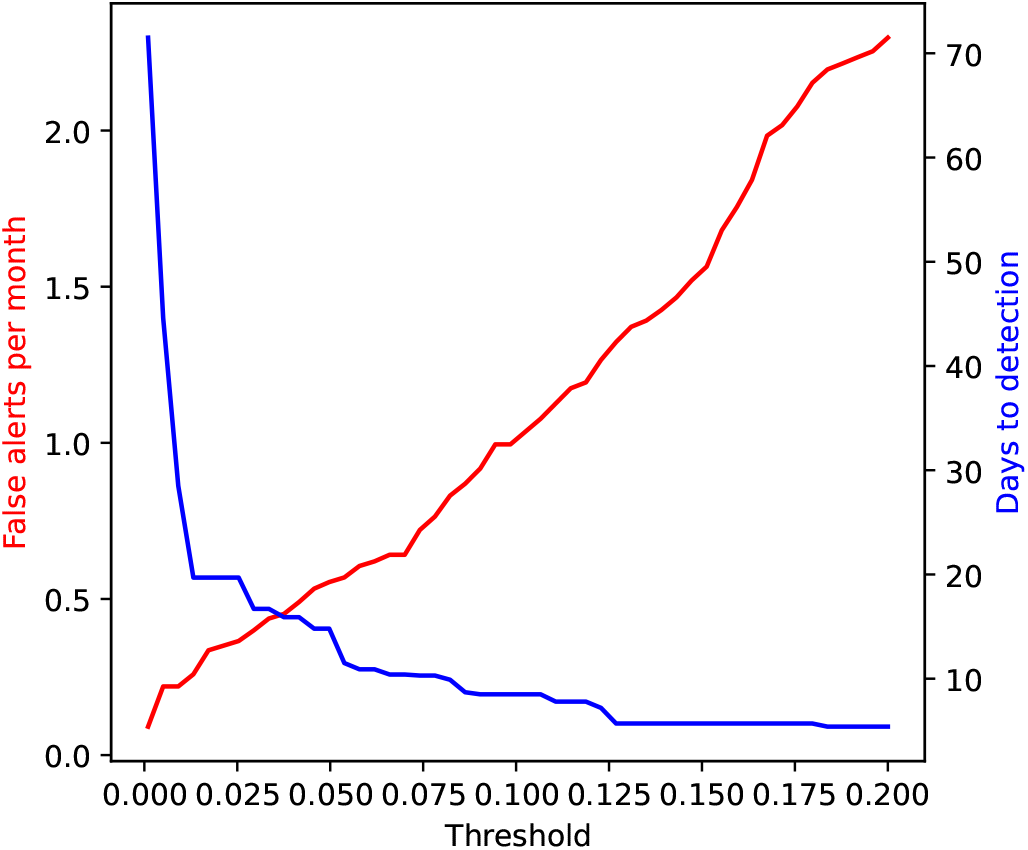
False alerts per month and days to detection.

An *activity monitoring operator characteristic* (AMOC) curve is a graph that characterizes the timeliness of a detector [15]. It plots the expected time to detection as a function of the false-positive rate. We can visualize the same information using the AMOC curve in Figure 7 that compares the number of false alerts to detection time.

**Figure 7.**
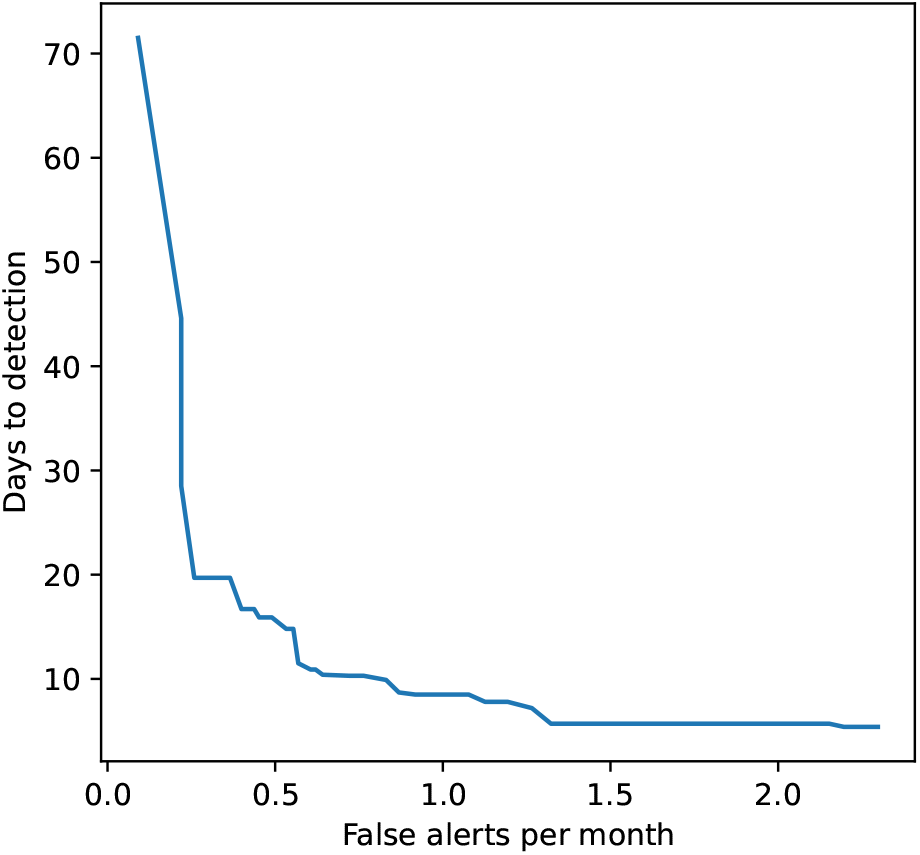
An average of the AMOC curves for detecting RSV and influenza during the years 2016-2021.

## Discussion

We applied ILI Tracker to a large, real-world dataset of records from UPMC EDs from 2011 through 2021. It was able to track multiple, often simultaneous, outbreaks of several ILIs using a Bayesian filtering approach.

There was a strong correlation of the ILI disease predictions with labeled influenza and RSV cases across multiple seasons. However, there was low correlation with the other five modeled ILIs. This result may be due to several factors. First, there are more labeled influenza and RSV cases, supporting better modeling and tracking. Also, RSV and influenza can be easier to diagnose since they are annual epidemics and when they circulate almost all other viruses do not appear.

In monitoring for the emergence of COVID-19, ILI Tracker identified significant drops in *p*-values as early as January 2020, well before laboratory confirmation in the study EDs. Additional low *p*-values throughout 2020 may reflect the arrival of new viral strains with altered clinical presentations. These findings support that the system was able to provide an early signal of unusual case patterns that are consistent with a novel outbreak prior to formal diagnostic recognition.

The artificial outbreak experiments further evaluated ILI Tracker’s ability to detect a novel outbreak. When confirmed COVID-19 cases were synthetically injected into historical ED data, the system produced a marked drop in *p*-values, consistently falling below 0·01 within two days. This result provides support that ILI Tracker could have detected a sudden appearance of COVID-19 cases, even if they had occurred in a different time period than early 2020.

The leave-one-out experiments, in which models for influenza or RSV were intentionally omitted, further evaluated ILI Tracker’s capability to flag unmodeled diseases. Across ten such tests, the system detected the omitted outbreaks with varying timeliness depending on the alert threshold, revealing a clear trade-off between rapid detection and false alert rates. When allowing on average of one false alert per month, the system is able to detect outbreaks in approximately eight days.

Empirical *p*-values generated by ILI Tracker—a measure of the unusualness of data—showed significant correlation to the start of the COVID-19 outbreak in early 2020, even as early as January of that year. This result, and additional experiments with injected data, support that ILI Tracker can detect the presence of a novel, unmodeled disease.

The ILI Tracker system has several advantages. It uses real-time data that are routinely collected in EDs. It is based on a principled Bayesian methodology that is transparent and explainable. In addition, it can readily incorporate additional disease models. Finally, although not emphasized in this paper, ILI Tracker is computationally efficient in handling large sets of findings, patients, and disease models [1].

ILI Tracker has some limitations. It relies on accurate NLP feature extraction and lab data. This can result in reduced accuracy in tracking diseases with low prevalence or subtle presentation. It assumes that diseases are stationary across seasons, so models trained on prior years may not be accurate if diseases change significantly. It also models the likelihood of findings given a disease using a Naïve Bayes approximation; this assumption could be relaxed in the future by modeling diseases using Bayesian networks, for example. The disease models of ILI Tracker do not incorporate epidemiological dynamics of the diseases, although this is also an area for future research.

Additional work should focus on modeling and distinguishing ILIs. A fundamental difficulty of modeling ILIs is that many ILIs have similar symptoms. Also, some non-ILI diseases that we classify as other—such as asthma—often have symptoms common to ILIs and may be (probabilistically) classified as an ILI. Explicit modeling of different diseases now included in *other* may improve performance.

ILI Tracker is available for use in public health practice [16]. Real-time deployment in EDs and other high-volume healthcare settings could help clinicians and public health officials recognize the emergence of new disease outbreaks earlier.

## Data Availability

The source data used in this project are protected health information and therefore cannot be shared externally.

## Acknowledgements

This research was supported by grant R01LM013509 (Automated Surveillance of Overlapping Outbreaks and New Outbreak Diseases) from the National Library of Medicine (NLM) of the U.S. National Institutes of Health (NIH). Harry Hochheiser and Jessi Espino also received support from NIGMS grant U24GM132013 (MIDAS Coordination Center) and NIGMS grant R24GM153920 (MIDAS Coordination Center). Ye Ye also received support from NLM grant R00LM013383 (Transfer Learning to Improve the Re-usability of Computable Biomedical Knowledge). Marian Michaels also received support from CDC grant U01IP001152 (New Vaccine Surveillance Network). Jessi Espino also received support from CDC grant 5U01IP001184 (Evaluating Respiratory Virus Vaccine Effectiveness in a Large, Diverse Healthcare System).

## Author Contributions

All authors contributed to the writing of this paper. John Aronis provided conceptual formulation, mathematical modeling, implementation and testing of the ILI Tracker system. Ye Ye provided conceptual formulation, mathematical modeling, implementation and testing of patient modeling. Jessi Espino provided conceptual formulation, data management, and extraction of MetaMap findings. Marian Michaels provided conceptual formulation, and clinical expertise. Harry Hochheiser provided conceptual formulation. Gregory Cooper provided conceptual formulation, clinical expertise, and mathematical modeling.

## Ethics Declarations

The research protocol was approved by University of Pittsburgh IRB Study 20030193.

## Data and Code Availability

Source code for ILI Tracker is available at https://github.com/RodsLaboratory/PDS. The source ED data used in this project are protected health information and therefore cannot be shared externally.

